# It’s complicated: characterizing the time-varying relationship between cell phone mobility and COVID-19 spread in the US

**DOI:** 10.1101/2021.04.24.21255827

**Authors:** Sean Jewell, Joseph Futoma, Lauren Hannah, Andrew C. Miller, Nicholas J. Foti, Emily B. Fox

## Abstract

Restricting in-person interactions is an important technique for limiting the spread of severe acute respiratory syndrome coronavirus-2 (SARS-CoV-2). Although early research found strong associations between cell phone mobility and infection spread during the initial outbreaks in the United States, it is unclear whether this relationship persists across locations and time. We propose an interpretable statistical model to identify spatiotemporal variation in the association between mobility and infection rates. Using one year of US county-level data, we found that sharp drops in mobility often coincided with declining infection rates in the most populous counties in spring 2020. However, the association varied considerably in other locations and across time. Our findings are sensitive to model flexibility, as more restrictive models average over local effects and mask much of the spatiotemporal variation. We conclude that mobility does not appear to be a reliable leading indicator of infection rates, which may have important policy implications.

## 1 Introduction

In the hopes of better informing public health decision-making, researchers have developed many prediction models to forecast the COVID-19 pandemic. Effective forecasts capable of identifying reliable leading indicators of emerging outbreaks could improve policy recommendations. To this end, factors such as mask-wearing [1; 2], weather [3; 4], and demography [5] have been found to be associated with rates of infection in the US. The effectiveness of other non-pharmaceutical interventions (NPIs) such as government lockdowns is also well studied [6; 7; 8], although some questions still remain. For instance, it is challenging to disentangle the effects of overlapping NPIs, such as the rapid increase in mask-wearing in early April 2020 alongside widespread lockdowns in many parts of the US.

Cell phone mobility data has emerged as an appealing surrogate of government mandates. Since it is a directly observable measure of human movement, it contains more information than the duration of government orders. In addition, it may serve as a better proxy for the actual quantity that government actions are intended to reduce: the relative frequency of risky in-person interactions where transmissions may occur. Mobility information is available through public APIs such as Google’s Community Mobility Reports [9] and SafeGraph’s completely at home metric [10]. The ubiquity of readily accessible mobility data, along with the lack of widespread alternative sources of data—such as detailed contact tracing information—has made it an attractive proxy for interactions.

As mobility plummeted to unprecedented levels during the first wave of the pandemic, these publicly available data sources received widespread attention. Mainstream media outlets such as the Washington Post [11; 12], Wall Street Journal [13], New York Times [14], Los Angeles Times [15], and National Public Radio [16] have all analyzed cell phone mobility and highlighted its record drop over the past year. Moreover, public-facing epidemiology dashboards, such as the ones available by the US Centers for Disease Control and Prevention [17] and the Institute for Health Metrics and Evaluation [18], prominently list mobility as a metric of interest. As articles in leading scientific journals began to suggest that mobility data could be a valuable tool for battling the pandemic [19; 20; 21], it is not surprising that many COVID-19 forecasts have used mobility as a data source.

Although there is a large and growing body of work using mobility to predict COVID-19 spread, many of their conclusions are not broadly applicable outside of the initial wave of the pandemic. In particular, limitations in the amount of data and in the inherent modeling assumptions restrict the applicability of these earlier works [22; 21; 8; 23; 24; 25]. Since the pandemic has been constantly evolving, a rather obvious limitation is that early papers only looked at data from the first few months of the pandemic, such as through June 2020 [22; 21; 8; 23; 24]. Furthermore, most articles limited the set of locations modeled to a small number of major cities [22; 21], or fit models at a coarser state level [8; 25]. Such limitations in the length of time and number of locations modeled render these works incapable of making inferences about local outbreaks across time. Another key limitation in most prior work—with the exception of [24]—is the overly restrictive assumption that the relationship between mobility and infection rates is stationary. Although this stationarity assumption was reasonable during the initial wave of the pandemic, large shifts in behavior due to evolving government guidance and adherence to such guidance suggest that coarse mobility may no longer be a good proxy for potentially risky transmission events [26; 27]; as such, the relationship between mobility and infection rates today likely differs from spring 2020.

Capturing the time-varying relationship between mobility and infection rates is especially challenging due to the incomplete, heterogeneous, and non-stationary nature of the data. For instance, the lack of reliable data on adherence to mask-wearing during the beginning of the pandemic in spring 2020 makes it difficult to identify the relationship between mask-wearing and infection rates. This problem is exacerbated by the fact that it is important to adjust for mask-wearing when interpreting the effect of mobility on growth rates. Reported case data come with their own set of unique challenges, including highly variable reporting delays, strong day-of-week effects, and differential rates of testing. Moreover, since we only observe this data over a relatively short time frame, it difficult to adjust for seasonality.

To assess the temporal and spatial utility of mobility data in this challenging data setting, a central objective of this work is to identify a flexible yet interpretable class of models that can sufficiently disentangle how the effect of mobility changes over time and space. To this end, we show that restrictive models effectively average over local effects by naively assuming a constant relationship between mobility and transmission. Conversely, we show that overly flexible models lead to spurious correlations and conclusions.

Our proposed multilevel regression model strikes a balance: we allow the association between mobility and growth rates to vary across groups of nearby counties and over four distinct “waves” of 13 weeks each. The granularity of this spatial clustering and temporal variation of coefficients is critical to the robustness of our inferences. We analyze an entire year of data across 94% of all 3,143 US counties (covering 99.7% of the total population) and use Google’s Mobility Trends as our measure of mobility. We replicate prior work that found strong first wave associations between mobility and infection rates. Furthermore, we find that the strength of this association is strongest in the most populous counties, but is otherwise highly variable across geographies, and significantly weakens after the first wave.

## 2 Results

### Visualization of mobility measures and infection growth rates over time

We first examine weekly county-level mobility and infection growth rate trends. Figure 1 visualizes the weekly growth rate of new infections for each of the 2, 951 US counties modeled. Counties are displayed according to the nine US Census divisions—within a division, counties falling in the same combined statistical area (CSA—a grouping of counties connected by workplaces and commuting patterns) appear in adjacent rows. Counties in the same CSA tend to exhibit similar growth rates, as evidenced by the clear clustering patterns in growth rates. Different waves of the pandemic across divisions are also apparent.

**Figure 1:**
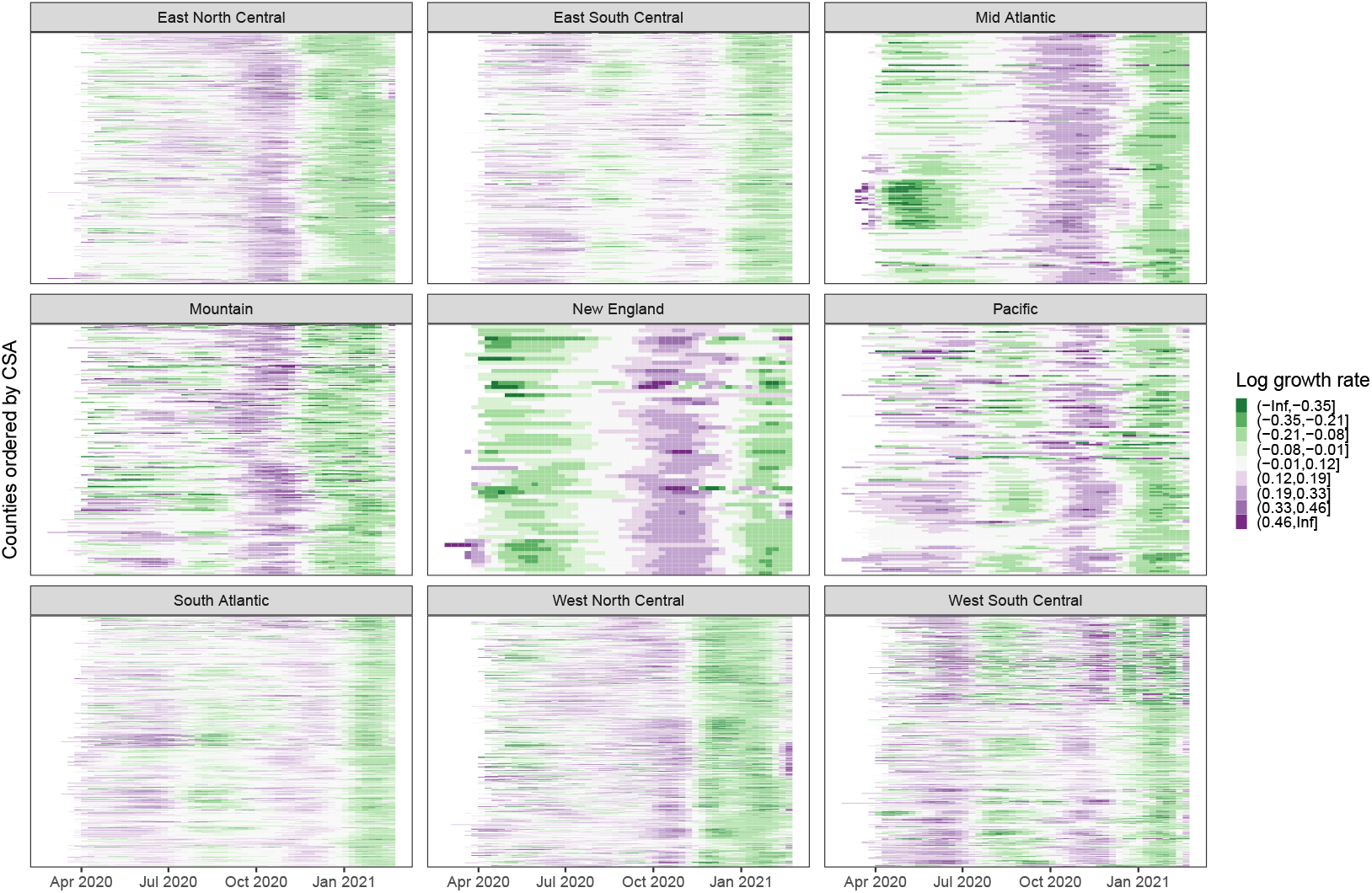
Weekly log growth growth *y*_*i,t*_ for all counties *i* organized by US Census division and CSA. Rapidly declining growth rates in New York City display prominently in the Middle Atlantic division in April 2020. The national surge in fall 2020, followed by declining infection rates in early 2021 is also pronounced.

Google’s mobility trends capture six distinct types of mobility: grocery/pharmacy, residential, retail/recreation, workplace, transit, and parks. Figure 2 shows the weekly trend for each of these variables for each county in three CSAs: New York City, San Francisco, and Green Bay, WI. Mobility values are reported relative to a baseline level in January 2020 for each county, which normalizes for population and pre-pandemic mobility levels. The rapid drop in mobility following widespread lockdowns in March 2020 is present in all locations. Furthermore, it is clear that these six mobility variables are tightly connected: grocery/pharmacy, retail/recreation, workplace, and transit are positively correlated, while residential mobility is negatively correlated with the others.

**Figure 2:**
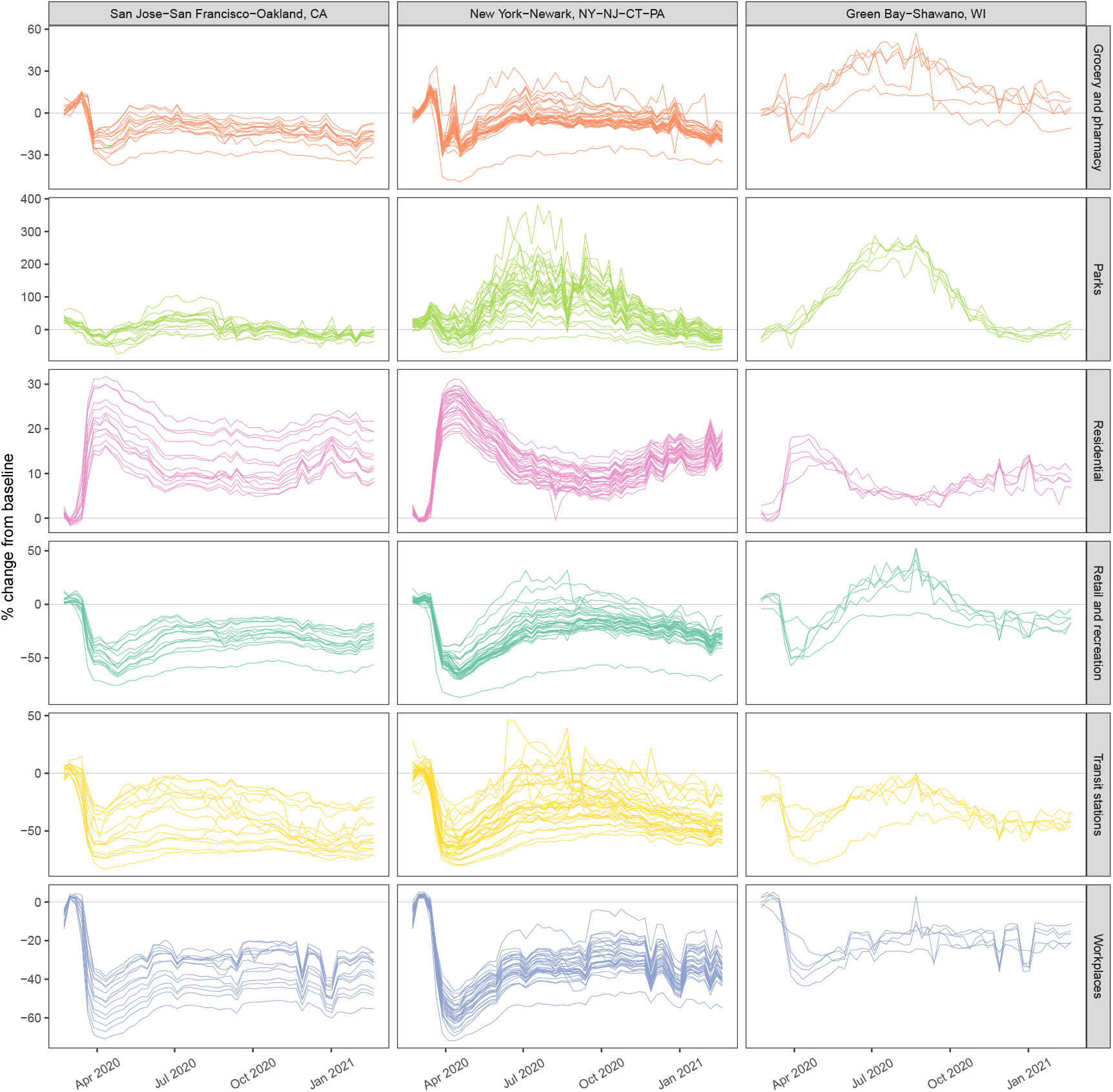
County-level weekly % change from baseline mobility for six mobility categories (grocery and pharmacy, parks, residential, retail and recreation, transit stations, and work-place) in three CSAs.

### Overly flexible models lead to incorrect and misleading inferences

It is tempting to include each of the Google mobility variables as separate predictors. However, the strong correlations between them often lead to misleading estimated associations between distinct mobility variables and growth rates. The “collinear” column of Figure 3 illustrates what can go wrong, showing two highly correlated mobility measures (retail/recreation and workplace) over time in one CSA. Nonsensically, the learned association between retail mobility and infection rates is negative throughout the first three waves. This misleadingly suggests that higher levels of retail-related mobility correlate with lower infection rates, but is clearly an artifact of collinearity between retail and workplace mobility. In our final model, we collapse the original six mobility measures into a single value using principal components analysis to avoid such unintentional side effects caused by collinearity [28]. This univariate feature captures over 60% of the variability in the original six mobility measures.

**Figure 3:**
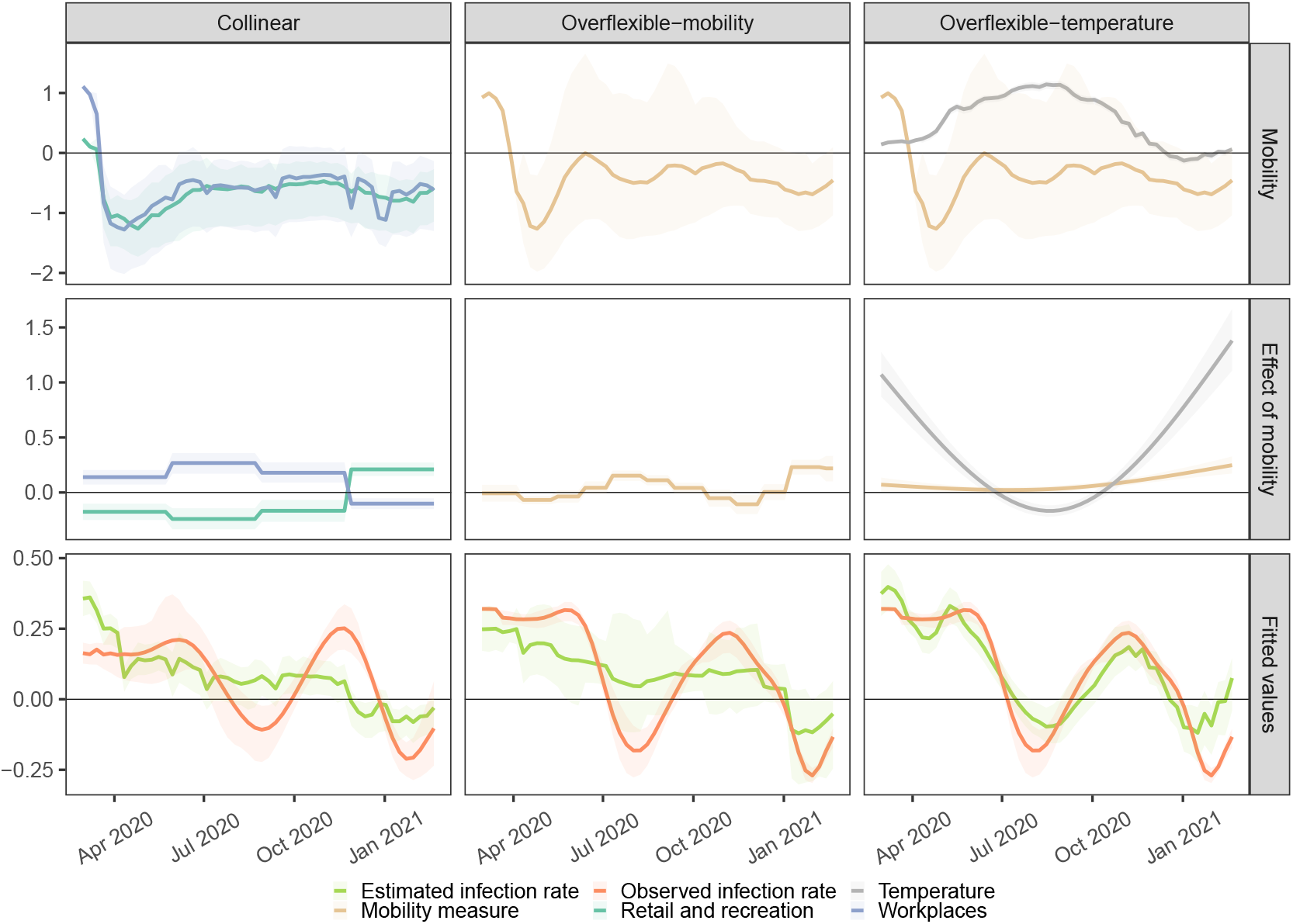
Illustrative model shortcomings due to collinearity in covariates (left), too much flexibility in mobility (middle), too much flexibility by including temperature (right). Observed covariate values (top), estimated effects of mobility (middle), and fitted and observed growth rates (bottom) are shown. Median (solid lines) and 95% quantiles (shaded) are shown.

The “overflexible-mobility” column of Figure 3 identifies another pitfall associated with too much model flexibility. The first principal component of mobility is visualized, along with its estimated association with growth rates from an overly flexible model that allows for the association to vary each month. The model’s effect of mobility over time for this location varies considerably, and often is negative. However, it is implausible that increased levels of mobility might be associated with lower infection rates; at most, there may be periods where they are uncorrelated.

The “overflexible-temperature” column Figure 3 shows results from allowing both the effect of temperature and the univariate measure of mobility to vary smoothly over time. Allowing the effect of temperature to also vary over time overpowers much of the signal contained in mobility, and it is clear that the model is over-specified by the near-perfect fit observed.

### Properly constrained models lead to meaningful inferences

As a first set of qualitative checks, we display in Figure 4 how well the model fits the observed infection rates for three CSAs chosen to illustrate heterogeneity in conclusions and model fit. Each column shows results from San Francisco, New York City, and Green Bay, WI. For each location, the aggregate mobility metric is plotted over time, along with the model’s coefficients for mobility per wave and the fitted and observed infection rate values. New York had a strong association between mobility and growth rates in the beginning of the pandemic, Green Bay had a strong association later in the pandemic, and San Francisco never had a strong association.

**Figure 4:**
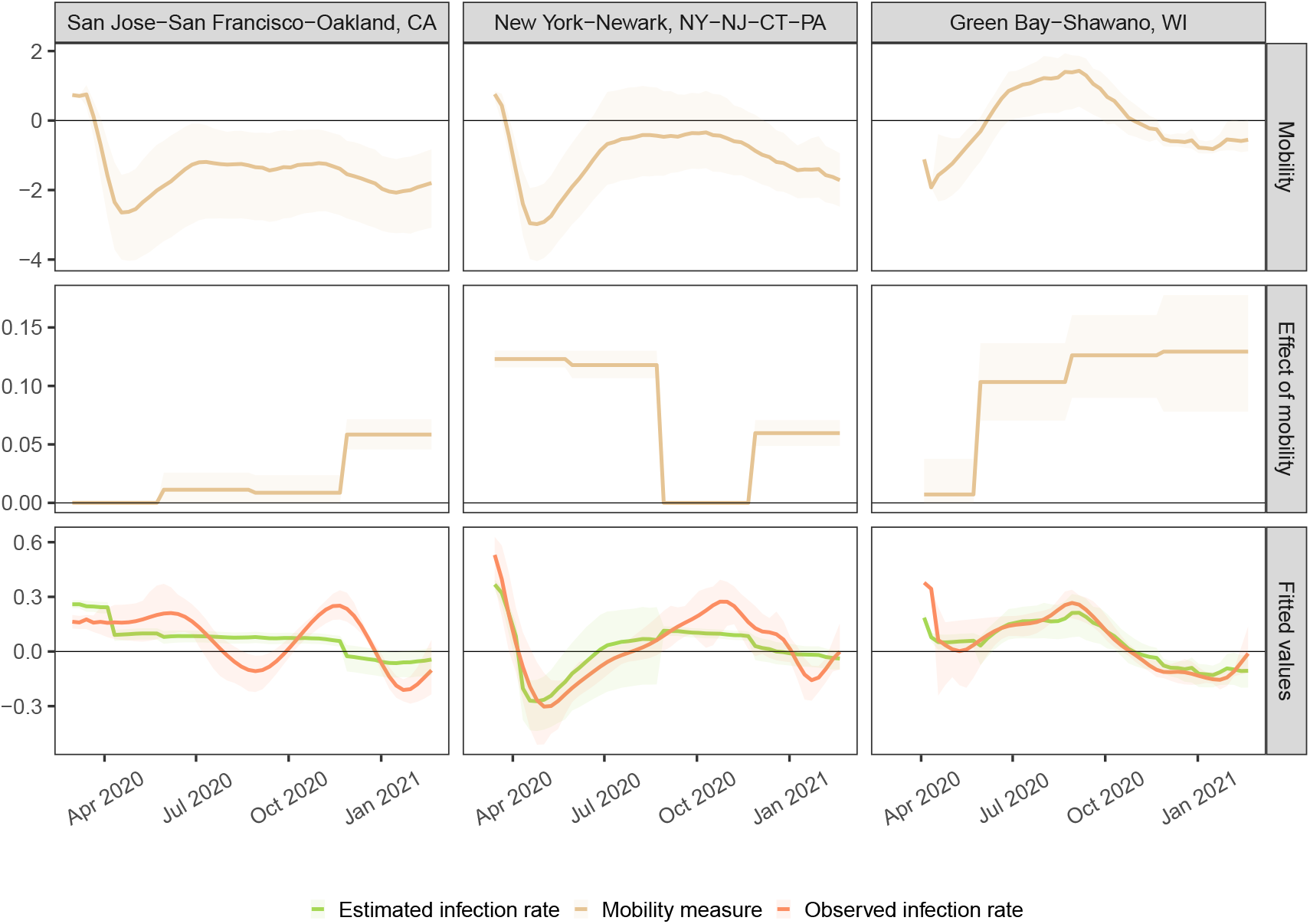
Mobility (top), estimated effect of mobility (middle), and fitted and observed infection growth rate values (bottom) for three illustrative CSAs. New York has a strong estimated effect of mobility in waves one and two, whereas Green Bay has a strong estimated effect of mobility in the second to forth waves. San Jose has a moderate effect of mobility in the fourth wave. Median (solid lines) and 95% quantiles (shaded) are shown.

### Mobility was most predictive in urban areas during spring 2020; elsewhere exhibited substantial variation

Figure 5 presents the *R*^2^ of our model across different subsets of data. Panel (a) shows the overall *R*^2^ of the model for each week and the *R*^2^ across counties with varying population sizes. The overall fit is best during the first months of the pandemic and for the largest counties (populations of more than 250, 000, comprising 64% of the total US population). *R*^2^ is low across the rural 46% of counties with a population of less than 25, 000. Panel (b) shows similar *R*^2^ results according to US Census region. The Northeast exhibits the best fit while the South, with its many rural counties, is the worst. Panels (c) and (d) show additional *R*^2^ results as a function of overall relative mobility levels across all locations and time. Model performance is highest during the first wave in the most urban counties, when mobility levels are at their lowest values. Interestingly, during the third and fourth waves there is minimal difference in *R*^2^ as a function of mobility levels, suggesting that at this coarse level of analysis mobility’s association with infection growth rates weakened over time.

**Figure 5:**
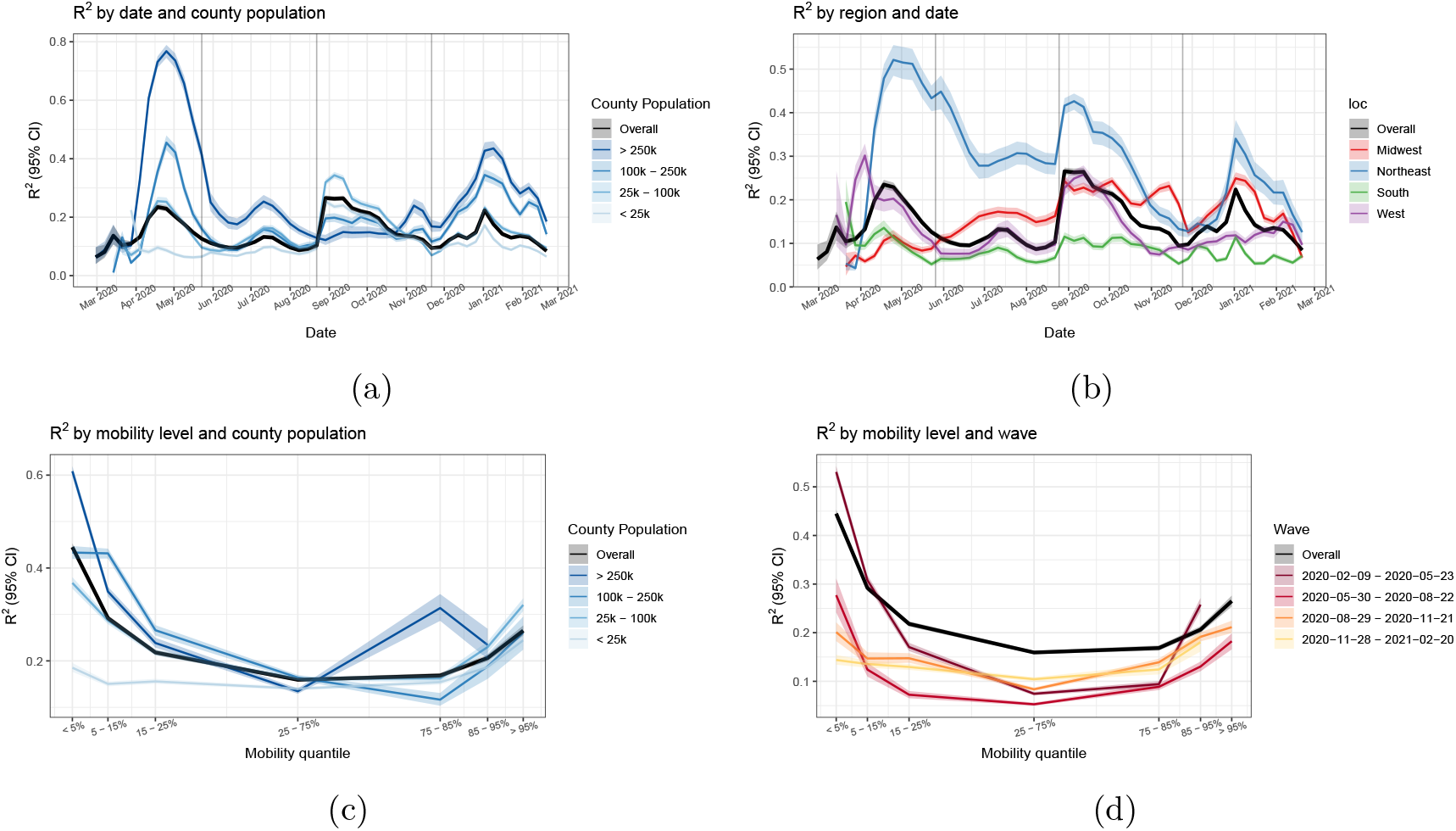
Model performance: (a) *R*^2^ per week, overall and by county population. (b) *R*^2^ per region and overall. At these coarse levels, models fit the best during April - May 2020.Fits were poor in summer, and improved in some places during fall and winter, but never return to the initial high levels. (c) *R*^2^ as a function of the overall level of mobility, further broken down by county population. Median (solid lines) and 95% quantiles (shaded) are shown. Mobility contains the most signal in the highest population counties when its overall value is extremely low. (d) *R*^2^ as a function of the overall level of mobility, further broken down by wave. Mobility contains the most signal in the first wave at extremely low values.

In Figure 6 we visualize the effect of mobility alongside the corresponding *R*^2^ for each wave and CSA on a map of the US. There is a striking degree of non-stationarity in the estimated effects over time and space. In the first wave, the estimated effect of mobility is close to zero throughout most of the South, as well as much of the West and Midwest. The signal weakens considerably in the second wave, while in the third wave the signal is strongest in the Midwest. Although the estimated effects of mobility sometimes appear strong, as in the fourth wave spanning winter 2020 into early 2021, the corresponding *R*^2^ values are often fairly weak.

**Figure 6:**
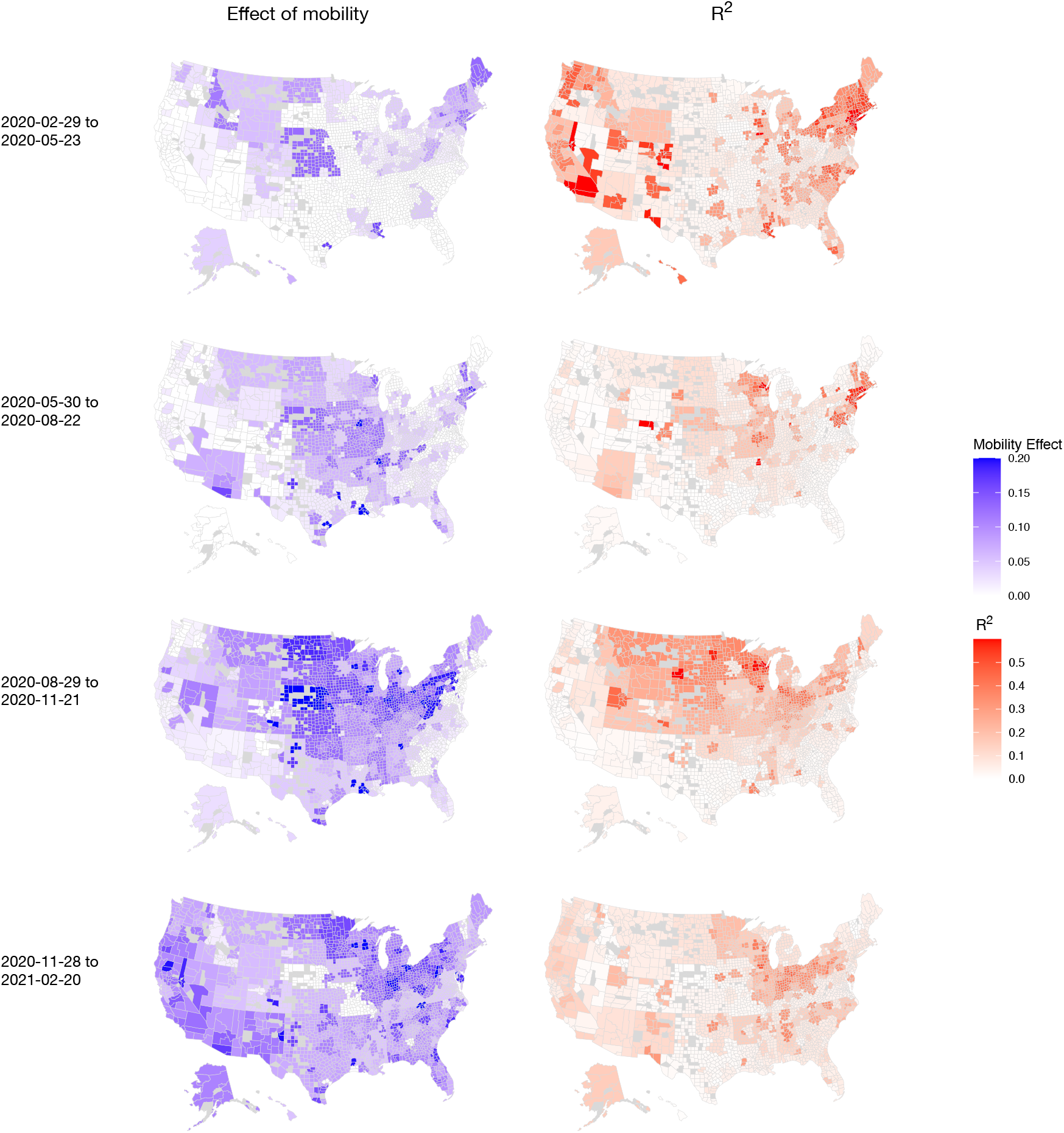
(Left) Estimated coefficients and (right) *R*^2^ by CSA across the 4 waves (rows).

### Overly rigid models underfit and wash out spatial and temporal effects

To assess whether our final model can be made simpler without sacrificing accuracy, we consider simpler models that limit mobility’s effect to vary by time and space. We construct an ablation study of six models: letting mobility’s effect vary by CSA, by region, or be fixed nationally; and letting mobility’s effect vary for each wave, or be fixed in time.

For the three example CSAs shown previously, we display the estimated effect of mobility across time for each ablation in Figure 7. Comparisons of models allowing differential effects of mobility across locations show that rigid national grouping averages over effects visible at finer spatial groupings, such as by region and CSA. Similar limitations are observed with constant temporal effects for mobility. This averaging is not just superficial: our conclusions on the association between mobility and the infection growth rate change. For example, in our final model, we conclude that there is no effect of mobility on infection growth rate in New York during the third wave. However, *all* other progressions would conclude that there *is* a strong association. Likewise, the simpler model that allows mobility’s effect to vary by CSA but forces it to be fixed in time would conclude that New York and Green Bay have very similar associations between mobility and infection rates. However, the final model clearly shows that they are actually quite different, as New York had the strongest association early on while the opposite trend held in Green Bay.

**Figure 7:**
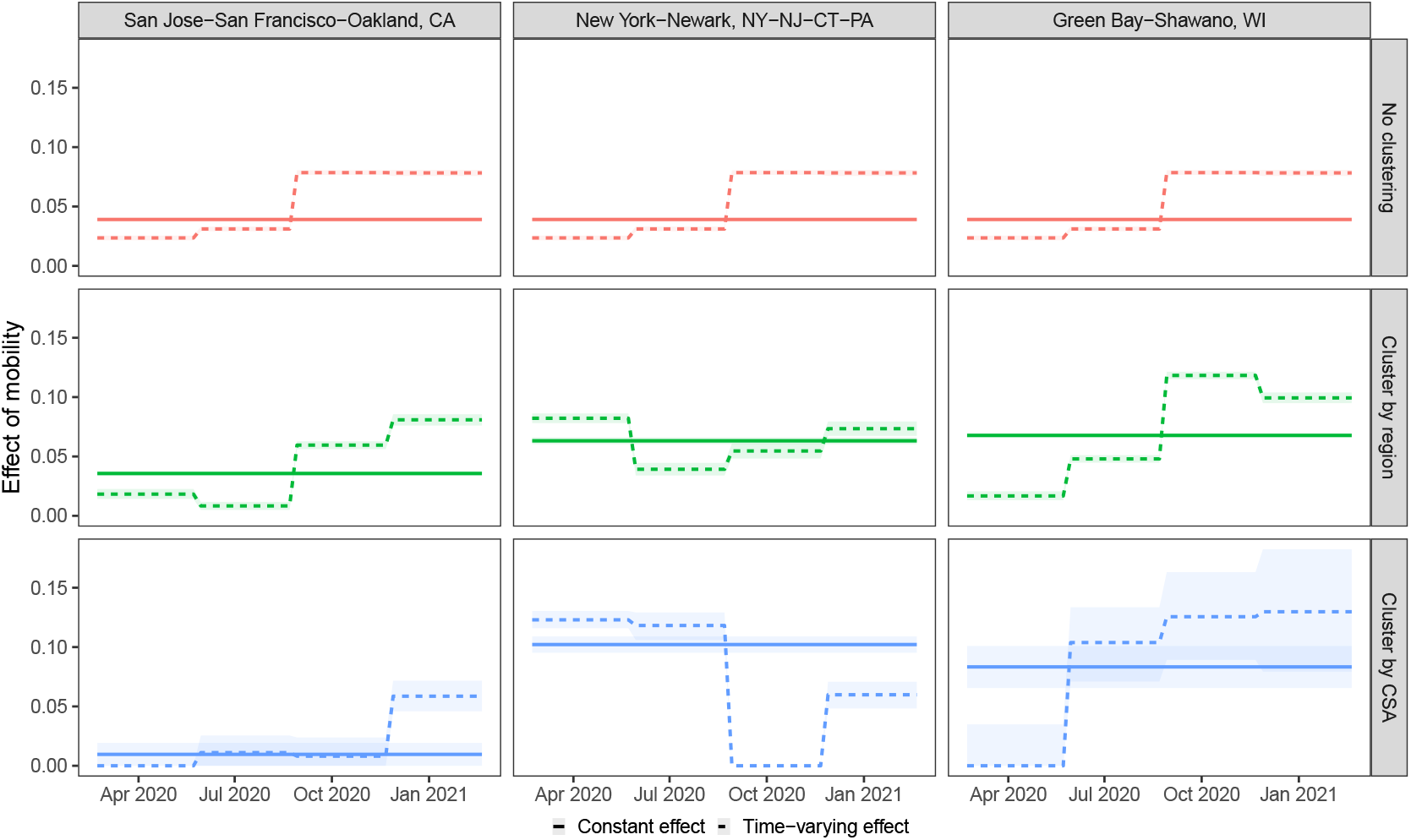
Overly rigid models average over spatiotemporal effects. The estimated effect of mobility for different spatial clustering (rows) and the form of temporal effects (line type) for three illustrative CSAs are displayed. Median (solid lines) and 95% quantiles (shaded) are shown.

Table 1 tabulates the overall and by region *R*^2^ for each of the six model progressions. As expected, greater flexibility generally results in higher overall *R*^2^. The greatest differences in *R*^2^ are observed at finer disaggregations: the simplest model has an *R*^2^ of just 19% in the North East, whereas our four wave CSA model achieves an *R*^2^ of over 40%; indicating that both time-varying coefficients and choice of clustering are critical. In Appendix C of the Supplementary Materials, we show that our final model does not overfit.

**Table 1:** *R*^2^ for no spatial clustering, clustering by region, or clustering by CSA, and constant or time-varying mobility coefficients.

| Clustering | Mobility Effect | Overall | Midwest | Northeast | South | West |
| --- | --- | --- | --- | --- | --- | --- |
| None | Constant | 0.163 | 0.201 | 0.186 | 0.123 | 0.175 |
| None | Time varying | 0.200 | 0.245 | 0.231 | 0.148 | 0.233 |
| Region | Constant | 0.188 | 0.275 | 0.279 | 0.108 | 0.158 |
| Region | Time varying | 0.216 | 0.318 | 0.289 | 0.125 | 0.196 |
| CSA | Constant | 0.204 | 0.291 | 0.303 | 0.121 | 0.175 |
| CSA | Time varying | **0.261** | **0.364** | **0.404** | **0.151** | **0.250** |

### Assessing the mask effect

On April 4, 2020, the Centers for Disease Control (CDC) began recommending public mask use, a stark reversal of earlier guidance. This led to an increase in mask use across the United States coincident with large drops in mobility. As a result of these concurrent events, mask use and mobility are strongly correlated in the first wave. To facilitate interpretation, we model the association between masks and the infection growth rate as a national effect that is constant across time. All other factors held constant, we estimate an expected 2% decrease in the infection growth rate due to an additive increase in mask adherence of 10%.

To untangle the effect of masks and mobility in the first wave, we compare the *R*^2^ by date in models with and without a mask variable. In the four week period following April 4, 2020, we find that overall *R*^2^ increases by approximately 10% when the mask variable is included in the model; see Supplementary Figure S3 for additional details.

### Conclusions are robust across different mobility data sources

To assess whether our conclusions are sensitive to choice of mobility measure, we consider SafeGraph’s completely at home data measure (`completely_home_prop_7dav`) [29; 10] in place of the first principal component of Google’s mobility indicators. Our conclusions are very similar when using either Google’s or SafeGraph’s mobility measure. Panel (a) of Figure 8 displays performance, as measured by *R*^2^, over time and by county population. As in Figure 5, *R*^2^ is highest in the beginning of the pandemic and in high population counties.

**Figure 8:**
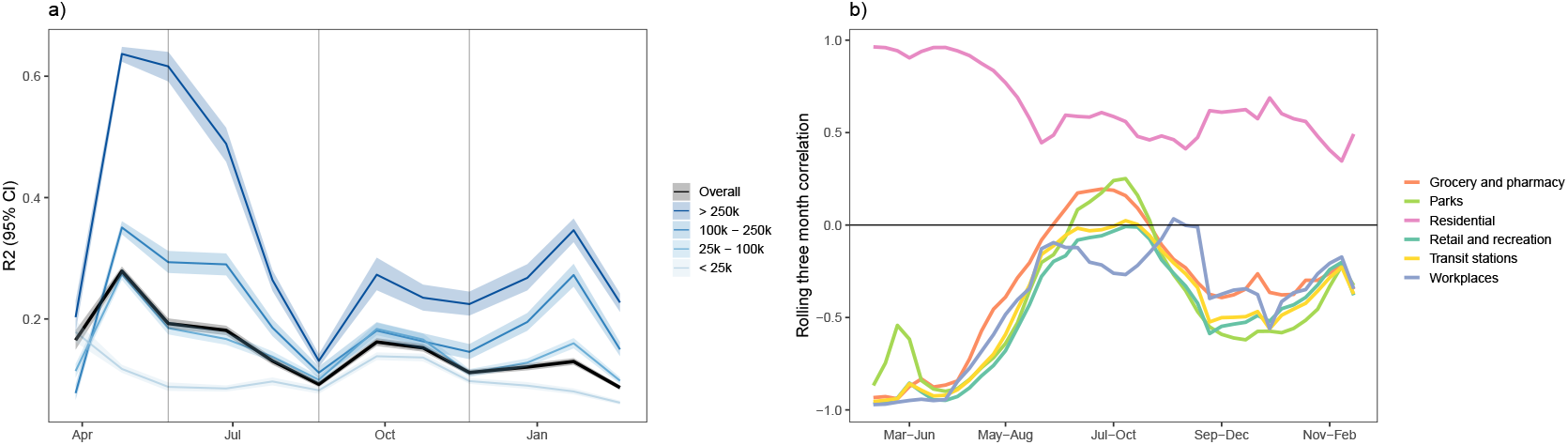
(a) *R*^2^ obtained using SafeGraph’s completely at home metric in place of the first principle component of Google’s mobility trends dataset. Median (solid lines) and 95% quantiles (shaded) are shown. (b) Rolling median three month correlation between SafeGraph’s completely at home mobility metric and each of the six Google mobility indicators. From March-June, absolute correlations were high, indicating consistency between all measures of mobility. However, the strong relationship decayed through May-October suggesting person-to-person contact patterns may not be well captured through coarse cell phone mobility after the initial period.

In panel (b) of Figure 8, the rolling three month correlation between SafeGraph’s completely at home measure and each of Google’s six mobility measures is plotted. From March–June 2020, we see correlations with large magnitudes across all variables, providing evidence that stay-at-home orders, lockdown orders, and general uncertainty resulted in a large correlated shift in mobility that is observable across different measures. As a result, during the first wave of the pandemic, *any* of these mobility measures should have similar ability to predict infection rates. However, as the pandemic progressed this relationship eroded, potentially suggesting that coarse cell phone measures of mobility began to capture different aspects of mobility and in ways that may not as reliably explain person-to-person contact patterns.

## 3 Discussion

The primary aim of our study is to disentangle how the association between mobility and COVID-19 infection rates varies across time and space. Our work is unique in that it fits time-varying models down to the county level for the vast majority of US counties with an entire year of COVID-19 data. This allows us to much more closely examine when and where broad claims do or do not hold, and to try to assess what drives those patterns.

We find that at an aggregate level, mobility was a strong predictor of COVID-19 weekly infection rates in the first wave, from February 29, 2020 through May 23, 2020. This is similar to findings in other studies, where cell phone mobility was lauded as a strong predictor in the US and globally during the early part of the pandemic [30; 31; 22; 32; 21; 24; 30]. A few later studies noted that mobility was markedly less effective as a predictor in the US after the first wave [26; 27; 33], which is supported by our findings. We found that the association between mobility and infection rates in the most populous areas largely diminished over the summer and into the fall, then briefly strengthened in late 2020 and into early 2021 before weakening again.

A complete understanding of the relationship between mobility and infection rates remains frustratingly elusive. Importantly, mobility is only a coarse proxy for a desired, but unmeasured quantity: the frequency of risky in-person interactions in a location, which should correlate more directly with infection rates. As in-person interactions changed over the last year due to better mask-wearing, hygiene, and social distancing, mobility data has become confounded and thus a worse proxy for risky interactions. We have also demonstrated that as mobility levels have slowly rebounded from extreme decreases seen during the first wave, different mobility measures have become less correlated. This implies that while *almost any* mobility metric would be a good proxy for risky interactions during extreme mobility decreases, much more care is required to select a proxy as mobility levels veer closer to pre-pandemic levels. We conclude that, while mobility was a reasonable proxy for less-safe practices at first, it was not necessarily stable through time or space.

In terms of modeling, our findings show that models that include mobility need to be either targeted to specific times and places, or include a relationship that varies with time and space. The latter is fundamentally challenging if other complicated relationships, such as with spatiotemporal-varying temperature or mask-usage associations, are included as well. The degrees of freedom quickly overwhelm data that is limited by collection period and correlations between explanatory variables. All statistical models used to understand relationships COVID-19 incidence and explanatory variables should be checked for the stability of coefficient values and predictive accuracy across time and space to avoid overfitting and spurious conclusions.

There are several limitations to these conclusions. From a data perspective, we face the fundamental problem of correcting for systematic differences in testing that occur over long periods of time and across locations. Although our use of growth rates provide some improvement over unadjusted case data, such period-by-period estimates cannot capture longer term trends in differential testing. While modeling hospitalizations could have addressed such issues, these data are not widely available at the county level. Another limitation arises due to a lack of detailed mask behavior during the initial phases of the pandemic, making the task of disentangling the effect of mobility and masks very difficult. Additionally, observed data is often systematically missing and must be imputed, and checking the embedded assumptions in our imputation models is challenging. Finally, we only observe a year of data which makes it impossible to correct for seasonality, such as with the effect of temperature. On a modeling side, we choose to pursue statistical models that estimate the association between mobility and the infection growth rate. Although these regression models do not allow us to simulate counterfactual scenarios as is possible with compartmental models [32; 21; 34; 35], such models are restrictive and subject to misspecification. To correctly specify these models would require knowledge of every infection for every county without reporting delays. Instead, infection count data is subject to changing protocols and availability, both of which are confounded by the dynamics of disease spread. In contrast, our regression framework is relatively easy to calibrate with existing data, and furthermore the simplicity of these models makes them much more computationally efficient to fit than compartmental models.

In terms of policy, our findings imply that public health officials should not focus exclusively on coarse mobility and must take into account other factors to measure possible transmission events. Conversely, our findings also suggest that there are settings where increased mobility does not necessarily indicate increased rates of transmission. However, the data are far too coarse to indicate what those settings are and what level and type of mobility would be safe. As states loosen mask usage and other restrictions, we might again see a changing effect of mobility. Furthermore, both the proliferation of more transmissible variants of the virus as well as the increasing number of vaccinated people will likely complicate the future relationship between mobility and COVID-19 transmission. These effects were not included in our analyses due to the time periods analyzed, but warrant future investigation.

## 4 Methods

### Overview

Our primary interest is to understand how the relationship between COVID-19 outbreaks and mobility varies across time and geography. Unfortunately, the exact time that new infections occur is never directly observed. Instead, we must rely on noisy observations of the infection incidence such as reported cases, hospitalizations, or deaths. To account for this discrepancy, we estimate the incidence of infections with a newly proposed statistical procedure that robustly estimates the true unknown infection incidence from reported cases [36]. We apply this estimator to daily reported cases for 2,951 counties (covering 99.7% of the total population) using data from the New York Times Coronavirus (COVID-19) repository [37] and the New York City Department of Health COVID-19 repository [38]; see Appendix A of the Supplemental Materials for data exclusion criteria.We then construct features from aggregated cell phone mobility data, mask-usage surveys, temperature data, and demographic data. These features are used to predict infection growth rates at the county level via a hierarchical Bayesian regression model.

### Infection growth rate as outcome

We hypothesized that mobility is more likely to correlate with the relative growth of an outbreak rather than with its absolute size. As such, we consider the log growth rate of the estimated incidence curve, henceforth referred to as the growth rate. Specifically, let *r*_*i,t*_ be the estimated number of new infections for county *i* that occurred in week *t*. As a unit-less quantity that measures the rate-of-change in the infection rate, the growth rate, *y*_*i,t*_, is more robust to differential testing rates than a quantity such as the estimated infection incidence itself. Define the weekly growth rate *y*_*i,t*_ as the log ratio of the total infections in the last two weeks: *y*_*i,t*_ = log (*r*_*i,t*_*/r*_*i*,t−1_).

Figure 1 displays the weekly growth rate of infections by geographic divisions, with counties ordered by their CSA. This figure illustrates the heterogeneity in the weekly growth rate: different regions experienced outbreaks of varying severity at different times. Similar temporal trends are observed not only within geographic divisions, but also in blocks of counties corresponding to CSAs. Our subsequent modeling choices that involve geographic hierarchies and wave break points are informed by this observed clustering.

### Data sources

We use Google’s publicly available mobility trends as a surrogate for the frequency of person-to-person contact. Google uses cell phone location data to measure the difference in movement trends during the COVID-19 pandemic from baseline activity before the pandemic for grocery/pharmacy, residential, retail/recreation, workplace, transit, and parks categories; see [9] for a detailed description. We apply weekly aggregation and imputation to account for day of week effects and missingness. To avoid collinearity in these features, as illustrated in Figure 3 of Section 2, we fit multilevel regression models with a univariate summary of mobility obtained as the first principal component of Google’s six mobility variables. For interpretation purposes, we enforce a positive first principal loading for workplace mobility, so that higher values of this summary variable indicate more time in public and less time at home.

State level mask usage features are created based on three surveys conducted at different times and at different geographic resolutions during the pandemic [39; 37; 29]. To easily interpret the expected effect of mask wearing, we combine responses from these surveys into a single continuous mask feature at the state level. Details are provided in Appendix A of the Supplementary Materials.

Mobility, mask use, and temperature time series are constructed by averaging daily measurements within each week. Google’s six mobility metrics and temperature are collected at the county level, and the mask feature is at the state level. The county level population is an estimate from the 2018 US Census.

### Multi-level regression model

We assume that the expected weekly infection growth rate in county *i* at week *t* is a linear function of population *X*_*i*_, temperature *T*_*i,t*_, mask compliance 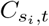, and a three week moving average of the first principal component of Google’s six mobility variables *M*_*i,t*_,

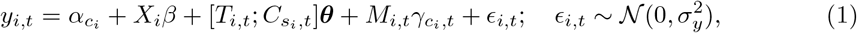

where *s*_*i*_ is the state of county *i*. To account for geographic clustering observed in the growth rate, we assume that the effect of mobility varies by CSA, i.e., 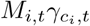, where *c*_*i*_ maps county *i* to its CSA^1^. This allows local information sharing—effectively augmenting missing or incomplete data—between counties within the same CSA. To account for non-stationarity in mobility, we assume that the effect of mobility on the infection rate varies across time. This is encoded through structured time-varying coefficients 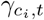.

For ease of interpretation, we assume the effect of mobility is piece-wise constant over four waves: February 22, 2020–May 23, 2020; May 30, 2020–August 22, 2020; August 29, 2020–November 21, 2020; November 28, 2020–February 20, 2021. This implies that 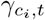 is a piece-wise constant function with three discontinuities and can thus be parameterized by four coefficients that describe the association between mobility and the infection rate in each wave. To prevent physically implausible coefficient values, we constrain the coefficients to be positive, that is, 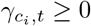 for all *c*_*i*_ and *t*. Markov chain Monte Carlo is used to sample from the posterior distribution. Appendix B of the Supplementary Materials contains a detailed description of our model, prior specifications, and posterior inference settings.

## Data Availability

We use publicly available data for county-level temperature, Covid-19 case counts, mask usage, Google mobility data, SafeGraph mobility data, and county population. SafeGraph mobility data and CMU mask usage surveys were openly available through the Delphi Epidata API (https://cmu-delphi.github.io/delphi-epidata/api/covidcast.html). All other data were openly available for direct download (https://goo.gle/covid-19-open-data, https://github.com/nytimes/covid-19-data, https://github.com/nychealth/coronavirus-data, https://github.com/nytimes/covid-19-data/tree/master/mask-use, https://www.pewresearch.org/fact-tank/2020/06/23/most-americans-say-they-regularly-wore-a-mask-in-stores-in-the-past-month-fewer-see-others-doing-it, https://www.google.com/covid19/mobility/, https://github.com/JieYingWu/COVID-19USCountylevelSummaries/tree/master/data).

https://github.com/nytimes/covid-19-data

https://github.com/nychealth/coronavirus-data

https://github.com/nytimes/covid-19-data/tree/master/mask-use

https://www.pewresearch.org/fact-tank/2020/06/23/most-americans-say-they-regularly-wore-a-mask-in-stores-in-the-past-month-fewer-see-others-doing-it

https://www.google.com/covid19/mobility/

https://github.com/JieYingWu/COVID-19USCountylevelSummaries/tree/master/data

https://cmu-delphi.github.io/delphi-epidata/api/covidcast-signals/safegraph.html

https://cmu-delphi.github.io/delphi-epidata/api/covidcast-signals/fb-survey.html#behavior-indicators

## Data availability

We use publicly available data for county-level temperature [40], Covid-19 case counts [37; 38], mask usage [41; 39; 29], Google mobility data [9], SafeGraph mobility data [10], and county population [42].

## Code availability

Code to reproduce all data preprocessing, model fitting, and subsequent analyses will be made available on Github after review.

## Author Contributions

J.F. and S.J. are co-first authors to reflect equal contributions. J.F., S.J., L.H., A.M., N.F., and E.F. conceived of the study. J.F. and S.J. performed the data preprocessing, wrote the source code to perform the experiments, and analyzed the results. J.F. and S.J. wrote the first version of the paper. All authors revised the paper and approve the submission.

## Footnotes

1 A state pseudo-CSA is created for all counties within a state that do not belong to one of the 175 named CSAs.

