## Supplementary Materials for "It’s complicated: characterizing the time-varying relationship between cell phone mobility and COVID-19 spread in the US"

April 21, 2021

#### A Data

**Imputation** We impute missing temperature and mobility observations from Google's mobility trends using the Multivariate Imputation by Chained Equations (mice) R package [1]. The MICE algorithm imputes missing values by iteratively fitting a conditional distribution for each variable in a dataset and using it to fill in missing values. This procedure is repeated a number of times until convergence is achieved. We impute values using the predictive mean matching method in mice. We parameterize the conditional distribution for each variable as a linear model, conditioned on the other observed variables. We also allow for a temporal trend per variable (e.g. to allow there to be some trend for mobility) within each US Census division and within each CSA, parameterized by natural cubic splines. This allows each CSA and each division to have its own smoothly varying trend per variable. We fit 25 multiply imputed datasets, and take the mean of these imputations to use for our modeling.

**County exclusion criteria** We exclude counties with less than 250 total COVID-19 cases as of the last date considered, February 20, 2021, which removes 176 counties. Next, we exclude counties with extreme growth patterns, where any weekly absolute growth rate exceeds 2 (removing 8 counties), or absolute growth rates exceeds 1.5 and the county has less than $50,000$ people (removing 8 counties). These restrictions remove outliers that arise from difficult to model events, such as prison outbreaks in sparsely populated counties.

**Feature selection** In addition to mobility, mask adherence, temperature, and county population, we also considered adjusting for county level demographic, socioeconomic, and health related features. However, since these features are constant in time and our model includes a random intercept by CSA, these additional variables only account for intra-CSA variability. Empirically, inclusion of these variables did not improve performance and made interpretation more difficult. As a result, we excluded these features from our final model.

*

†These authors contributed equally

**Mask featurization** To construct a single measure of mask adherence over the course of the pandemic, we combine survey responses from a few different sources. Pew Research carried out two surveys on June 7, 2020 and August 8, 2020 and released aggregate survey responses at the division level [2], and the New York Times and Dynata ran county-level surveys from July 2, 2020–July 14, 2020 [3]. From September 8, 2020, CMU's Delphi Epidata group administered and reported state level daily mask adherence survey responses [4]. We use the COVIDcast Epidata R package to download mask survey responses from CMU's Delphi Epidata repository.

We define our mask adherence feature piecewise: Between the two Pew survey dates, we linearly interpolate such that the state mask value intersects the average survey response of all counties in a state from the New York Times survey on July 7. The slope of the interpolant is set to the trend between the state's corresponding June and August Pew division responses. From the value on August 8, we linearly interpolate to the CMU state level value on September 8. If this results in a decrease in mask adherence between August and September, we instead use a single interpolant from June 7–September 8 defined by two points: the average state level response from the New York Times survey on July 7, and the state level CMU value on September 8. This monotonicity constraint ensures that the mask adherence level does not increase too quickly between survey dates over the summer.

We further assume zero mask wearing from the start of the pandemic until one week after the CDC adjusted their mask wearing recommendation on April 4; prior to this date, the CDC recommended not wearing masks. From April 11 until June 7, the state mask value is equal to the June 7 value.

### B Detailed model description

We model the expected log growth rate in county $i$ at week $t \in \{1, \ldots, N\}$ as a linear function of log county population $X_i$, average $t$th week temperature $T_{i,t}$, mask compliance $C_{s_i,t}$ in county $i$'s state $s_i$, and the three week moving average of the first principal component of Google's six mobility variables (constrained such that workplace mobility's loading is positive) $M_{i,t}$ at week $t$ through a multilevel Bayesian regression model:

$$\begin{aligned} y_{i,t} &= \alpha_{c_i} + X_i\beta + [T_{i,t}; C_{s_i,t}]\boldsymbol{\theta} + M_{i,t}\gamma_{c_i,t} + \epsilon_{i,t} \\ \beta &\sim 1 \\ \boldsymbol{\theta} &\sim \mathbf{1} \\ \epsilon_{i,t} &\sim \mathcal{N}(0, \sigma_y^2), \end{aligned} \tag{B1}$$

where $c_i$ is a surjective mapping from county $i$ to its CSA, temperature and mask use are concatenated by column in the matrix $[T_{i,t}; C_{s_i,t}]$, the notation "$A \sim \mathbf{1}$" defines an improper flat prior over the reals for the random variable $A$. Log population estimates and weekly temperature observations are each centered by their mean and normalized by twice their sample standard deviation. $\gamma_{c_i,t}$ is chosen to have a specified parametric form that accounts for non-stationarity in the expected effect of mobility. We specify $\gamma_{c_i,t}$ through a fixed weight matrix $\boldsymbol{W} \in \mathbb{R}^{N \times R}$ and a cluster-specific vector $\boldsymbol{\rho}_{c_i}$ of dimension $R \ll N$ that parameterizes the coefficients

$$\boldsymbol{\gamma}_{c_i,t} = \boldsymbol{W}_t \boldsymbol{\rho}_{c_i},$$

where $\boldsymbol{W}_t$ is the $t$th row of the weight matrix $\boldsymbol{W}$. In practice, $N = 52$ as we model a full year of data.

In the piecewise constant model with $R = 4$ waves, $\boldsymbol{W}$ is specified with three fixed knot dates $d_1$, $d_2$, and $d_3$: the $t$th row of $\boldsymbol{W}$ is defined as $\boldsymbol{W}_t = \left[1_{(t \le d_1)}, 1_{(d_1 < t \le d_2)}, 1_{(d_2 < t \le d_3)}, 1_{(t > d_3)}\right]$. Here $1_A$ is an indicator function that equals one if $A$ holds, and is zero otherwise. Cluster $c_i$'s $\boldsymbol{\rho}$ coefficients are defined as $\boldsymbol{\rho}_{c_i} = \left[\rho^1_{c_i}, \rho^2_{c_i}, \rho^3_{c_i}, \rho^4_{c_i}\right]^\top$. In practice, we let $d_1$ be May 23, 2020, $d_2$ be August 22, 2020 and $d_3$ be November 28, 2020, as this evenly splits the 4 waves into groups of 13 weeks each.

We further specify a joint distribution over the coefficients $\alpha_{c_i}$ and $\boldsymbol{\rho}_{c_i}$,

$$\left[\alpha_{c_i}; \boldsymbol{\rho}_{c_i}^\top\right] \overset{\text{ind.}}{\sim} \mathcal{N}\left(\left[\alpha_0, \boldsymbol{\rho}_0^\top\right], \boldsymbol{\Sigma}\right),$$

where the covariance matrix $\boldsymbol{\Sigma}$ is defined through a scaled correlation matrix $\boldsymbol{\Omega}$ which is distributed according the LKJ distribution [5] with shape parameter equal to two,

$$\boldsymbol{\Sigma} = diag(\boldsymbol{\tau})\boldsymbol{\Omega} diag(\boldsymbol{\tau}).$$

The scales $\boldsymbol{\tau}$ are half t-distributed with three degrees of freedom. The population-level intercept $\alpha_0$ is $t$-distributed with three degrees of freedom and $\boldsymbol{\rho}_0 \sim 1$.

We enforce a post-hoc positivity constraint that $\gamma_{c_i,t} \ge 0$ by applying the function $f(x) = \max(0, x)$ to all samples from the posterior distribution of $\gamma_{c_i,t}$. We found that such post-thresholding generally led to similar estimates when compared to a model where the coefficients $\gamma_{c_i,t}$ were log-normally distributed (and thus satisfy $\gamma_{c_i,t} \ge 0$), but convergence and sampling time per MCMC iteration was much faster.

**Model training and evaluation** We use the R package brms to obtain posterior samples from model (B1). Two chains are run for 7000 total iterations; 2000 samples are used for calibration during warm-up. We set the adapt delta and max treedepth settings to 0.9995 and 25, respectively. Every fifth sample is retained for posterior inference. Final models are assessed to ensure convergence: all estimated $\hat{R}$ values are less than 1.05; tail and bulk effective sample sizes are all greater than 1000.

**Estimated coefficients from fitting model** (B1) Posterior estimates for population-level parameters are presented in Table 1. The mobility wave parameters (i.e. the $\rho_0$) can be interpreted as the mean effects over all CSAs; these point estimates are comparable to what the effects would be in a model that forces the same association across space and does not allow for differential effects by CSA.

### C Ablation studies

**Results are robust to choice of knot locations.** In Figure S1, we examine the sensitivity of our results to different knot locations. We consider 125 different models with differing knot dates. In our final model in the main paper, we let $d_1$ be May 23, 2020, $d_2$ be August 22, 2020 and $d_3$ be November 28, 2020, as this evenly splits the 4 waves into groups of 13 weeks each. We considered models where we jittered $d_1$ by up to 2 weeks before or after May 23, 2020 (i.e. we tested $d_1 \in \{2020-05-09, 2020-05-16, 2020-05-23, 2020-05-30, 2020-06-06\}$. Similarly, we jittered $d_2$ and $d_3$ by up to 2 weeks before and after their final dates as well, for a total of 125 different knot combinations.

For each model, we compute the overall $R^2$, $R^2$ by population, and $R^2$ by region. As shown in red the $R^2$ of our final model is roughly centered in each histogram; our final model is not overfit to knot locations.

| | Estimate | Lower 95% CI | Upper 95% CI | $\hat{R}$ | Bulk ESS | Tail ESS |
|---|---|---|---|---|---|---|
| Intercept ($\alpha_0$) | 0.19 | 0.18 | 0.20 | 1.00 | 1241 | 1530 |
| Population ($\beta$) | 0.04 | 0.03 | 0.04 | 1.00 | 1919 | 1776 |
| Temperature ($\boldsymbol{\theta}_1$) | 0.03 | 0.03 | 0.03 | 1.00 | 1882 | 1604 |
| Mask ($\boldsymbol{\theta}_2$) | -0.19 | -0.20 | -0.19 | 1.00 | 1978 | 1965 |
| Mobility Wave 1 ($\rho_0^1$) | 0.00 | 0.00 | 0.01 | 1.00 | 1123 | 1495 |
| Mobility Wave 2 ($\rho_0^2$) | 0.04 | 0.03 | 0.05 | 1.00 | 1132 | 1693 |
| Mobility Wave 3 ($\rho_0^3$) | 0.09 | 0.07 | 0.10 | 1.00 | 1101 | 1542 |
| Mobility Wave 4 ($\rho_0^4$) | 0.11 | 0.10 | 0.12 | 1.00 | 1703 | 1814 |
| Error scale ($\sigma_y$) | 0.13 | 0.13 | 0.13 | 1.00 | 1966 | 1965 |

Table 1: Posterior population level parameter estimates obtained from 2000 posterior samples (2000 for warm up, and 5000 remaining samples where thinned by saving every 5th sample) in each of two MCMC chains from (B1).

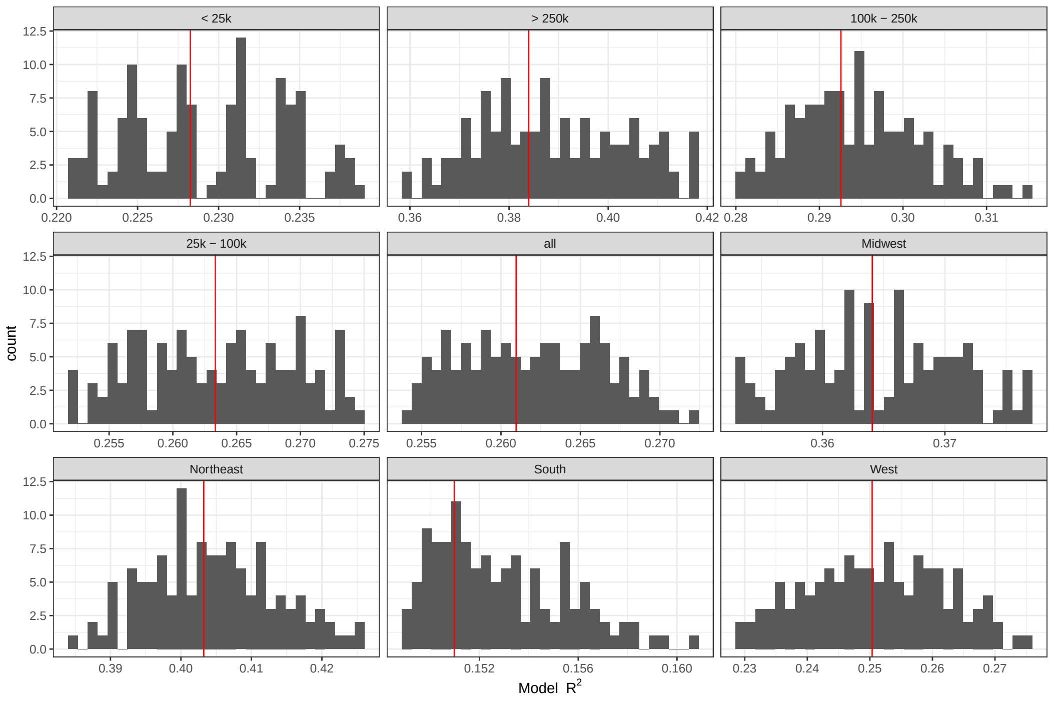


Figure S1: Distribution of $R^2$ values across 125 different models with slightly different locations for the knots defining the 4 waves. Each facet displays a histogram of $R^2$ across all models on that subset of data (either all of the data, one region, or counties of a certain population size). The red line in each pane shows our final model's $R^2$. Although performance could be slightly improved, quantitative performance is not very sensitive to the precise choice of knots.

**Averaging over many training and testing splits, our model does not overfit.** In order to confirm that our models are not overfit, we ran the final model 100 times each using two different strategies for constructing training and held-out testing sets. First, we created splits by holding out a random 20% of weeks ("random-times"), ignoring geography (i.e. some weeks in a given county will appear randomly in the training set, and some in testing). This specific type of split is less likely to exhibit overfitting, as there will generally be at least some data from every county. Second, we created splits by holding out all data from a random 20% of counties ("random-counties"), fitting the model on the remaining 80%. Figure S2 displays the $R^2$ for each data splitting strategy by week for train and test splits.

When averaging across 100 splits of randomly held-out times, the mean overall $R^2$ performance is 20.4% (95% CI: (20.2%, 20.6%)) in-sample and 20.3% (95% CI: (19.8%, 20.9%)) out-of-sample. Region-specific $R^2$ values are: Midwest, 29.1% (28.6%, 29.4%) in-sample, 29.0% (27.9%, 30.1%) out-of-sample; Northeast, 30.1% (29.5%, 30.8%) in-sample, 29.9% (27.7%, 31.9%) out-of-sample; South, 12.2% (12.0%, 12.4%) in-sample, 12.1% (11.5%, 12.6%) out-of-sample; West, 17.7% (17.3%, 18.2%) in-sample, 17.5% (16.5%, 18.6%) out-of-sample.

When averaging across 100 splits of randomly held-out counties, the mean overall $R^2$ performance is 26.2% (95% CI: (25.7%, 26.9%)) in-sample and 26.7% (95% CI: (24.9%, 29.2%)) out-of-sample. Region-specific $R^2$ values are: Midwest, 36.6% (35.7%, 37.5%) in-sample, 36.5% (33.2%, 40.9%) out-of-sample; Northeast, 40.7% (38.9%, 42.4%) in-sample, 40.4% (33.9%, 46.8%) out-of-sample; South, 15.2% (14.6%, 16.0%) in-sample, 15.7% (13.4%, 18.1%) out-of-sample; West, 25.4% (24.4%, 26.7%) in-sample, 25.4% (20.0%, 31.4%) out-of-sample.

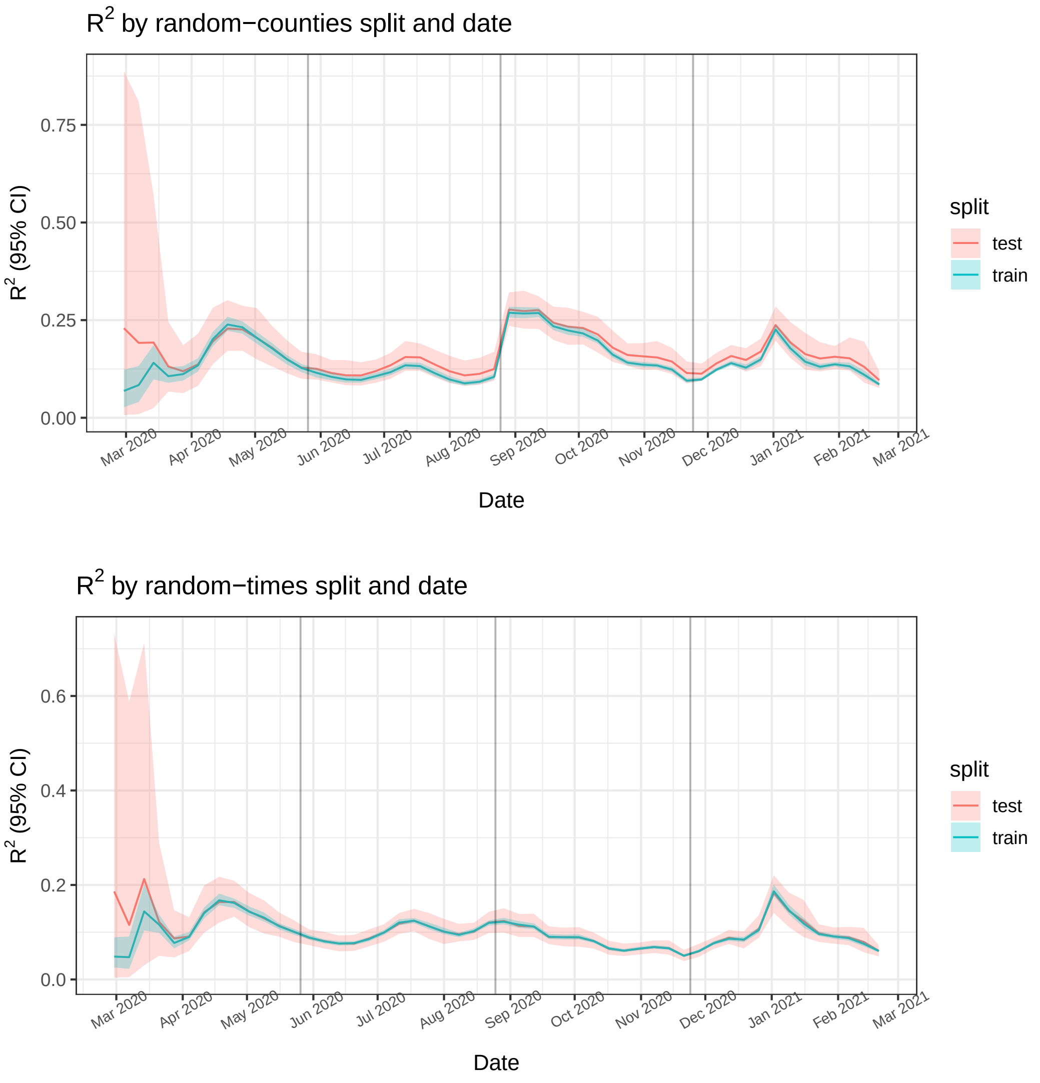


Figure S2: Train/test results, when disaggregating by week. Median (solid lines) and 95% quantiles (shaded) are shown.

**Adjusting for mask use leads to increased $R^2$.** To assess the effect of mask use, we compare two versions of our final model that differ only in whether or not they include or exclude the mask feature as a global fixed effect. Figure S3 shows that the model with masks included leads to substantial increases in $R^2$ in the first wave (approximately 10%) and moderate increases in the third wave (approximately 4%) over the model with masks excluded. Overall, the $R^2$ of 27.2% in the final model including masks is about 2% higher than the $R^2$ of 25.2% in the model excluding masks.

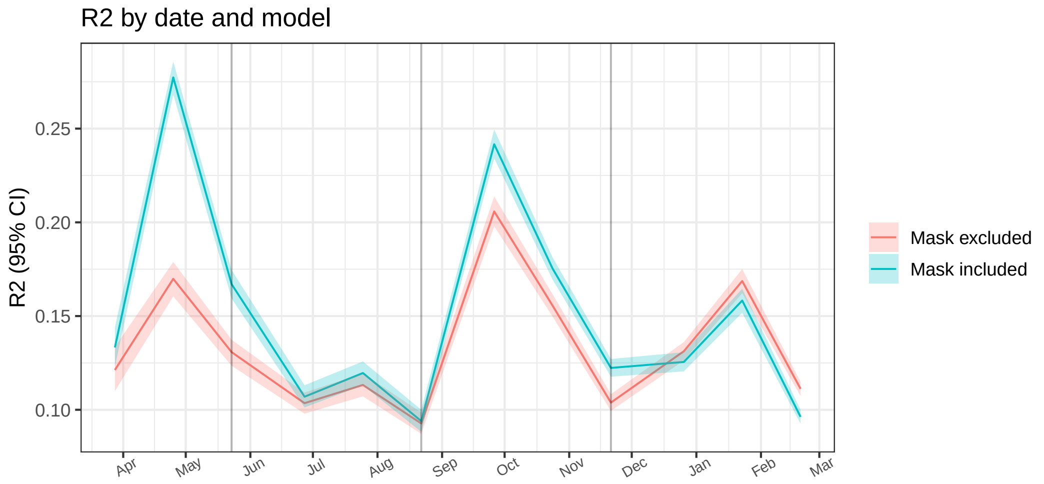


Figure S3: $R^2$ across time for base model with and without mask variable. Median (solid lines) and 95% quantiles (shaded) are shown.
